# Epidemiology of Extrapulmonary Tuberculosis in Brunei Darussalam: A retrospective cohort study

**DOI:** 10.1101/2022.11.21.22282609

**Authors:** Liling Chaw, Lena Mat Salleh, Rafizah Abdul Hamid, Kyaw Thu

## Abstract

**Objectives:** We reported the incidence and associated factors of extrapulmonary tuberculosis (EPTB).

**Design:** A retrospective cohort study.

**Setting:** Brunei Darussalam, an intermediate tuberculosis (TB)-burden country with stagnating annual TB rates.

**Participants:** All active TB cases identified in the country between January 2001 and December 2018 (18 years)

**Primary and secondary outcome measures:** Annual proportions of EPTB (overall and specific) were calculated. Multiple logistic regression was done to investigate factors associated with developing EPTB, when compared to pulmonary TB (PTB). Chi-square trend test was used to determine any trends during the 18-year study period.

**Results:** We identified 3,916 TB cases, among which 743 (19.0%) were EPTB cases. Lymphatic (44.8%) and pleural (19.4%) EPTB were most common. The main modes of diagnosis were tissue biopsy (73.6%) and radiologic assessment (18.3%). Treatment success and mortality rate were 79.7% and 7.0%, respectively. Associations with specific EPTB types varies with age-group and gender. Younger age-group (adjusted Odds Ratio [aOR] ≥1.94) and females (aOR: 2.45 [95%CI: 1.94,3.11]) had higher adjusted odds of developing lymphatic EPTB, but had lower adjusted odds of developing pleural EPTB [younger age-group (aOR ≤0.54) and female (aOR: 0.41 [95%CI: 0.17,0.90])]. When compared to foreign residents, locals had higher adjusted odds of skeletal (aOR: 4.44 [95%CI: 2.04,11.69]), gastrointestinal (aOR: 3.91 [95%CI: 1.84,9.66]) and other types of EPTB (aOR: 3.42 [95%CI: 1.53,9.14]). No significant trend differences were observed for overall and specific EPTB types.

**Conclusion:** Despite being generally non-infectious and less recognised than PTB, understanding EPTB epidemiology is important as it also contributes to the overall TB burden in a country. Examining EPTB cases by their specific anatomical site would provide more information on risk factors. Raising public awareness on the EPTB symptoms and that TB affects lungs and other parts of the body could promote early health seeking behaviour and early EPTB diagnosis.

**Strengths and limitations of this study:**

- - We analysed TB case notification data, retrospectively collected as part of the national TB surveillance programme.
- - The main study strength is the length of the study period (18 years) and that all notified TB cases in the country were captured in this dataset.
- - Study limitations include the inability to distinguish between PTB cases with only lung involvement and those with miliary TB and/or concurrent PTB and EPTB, and also the inability to include data on co-morbidities.

## Introduction

Tuberculosis (TB) is a bacterial disease that can affect both the lungs (defined as pulmonary TB [PTB]) and other body organs (defined as extrapulmonary TB [EPTB]). According to the World Health Organization (WHO), EPTB accounts for 16% of all notified 7.1 million incident TB cases in 2019 [1]. This proportion is likely an underestimate due to diagnostic challenges [2]. EPTB may present with symptoms which can be specific to the anatomic site affected and may mimic symptoms of other more common diseases. Also, diagnosing EPTB often requires site-specific microbiological evaluations which could be difficult to access depending on the affected site(s). Hence, national EPTB rates tend to vary depending on the socio-economic situation of the country, and also if adequate resources were allocated to their national TB programmes [2].

Recent epidemiological studies from other countries have reported increasing trends of EPTB prevalence, accompanied by decreasing trends of PTB prevalence [3–6]. The common anatomic sites of EPTB reported were lymphatic and pleural [3], while other studies also reported high incidence of urogenital, skeletal, gastrointestinal and meningeal EPTB [3,7]. Conflicting evidence exist on the risk factors associated with EPTB, using PTB as the comparison group. While risk factors such as being female, younger age, Asian and African ethnicity, and comorbidities such diabetes and HIV were found to be associated with EPTB [3,7–10], there were also other reports leaning towards being male [4,5] and increased age [4]. Although EPTB is generally less prevalent and also less infectious than PTB, it also contributes to the overall TB burden [2] particularly in terms of TB-related morbidity, complications and lifelong sequelae as a consequence of the disease [6]. This is thus important to understand the epidemiology of EPTB, particularly in settings with stagnating TB incidence.

Brunei Darussalam (pop. 440,715[11]) is a Southeast Asian country with intermediate TB burden, where the TB incidence was 61 per 100,000 population in 2021 [12]. Annual TB incidence for the country has remained stagnant since 2004 [13]. The incidence of both drug-resistant TB (0.3 per 100,000 population [12]) and HIV case counts (31 new cases in 2019 [14]) remained low. Non-communicable diseases are a major issue for Brunei, particularly diabetes mellitus whose prevalence was 11.1% among the adult population (20-79 years) in 2021 [15] and is also a known risk factor for TB [10]. Determining the factors associated with EPTB could assist in improving its early detection and/or diagnosis in the country, thereby allowing early treatment, prevent progression to infectious state and improving patient prognosis. Hence, we aim to determine the epidemiological characteristics of EPTB cases in Brunei and the factors associated with developing EPTB, when compared to PTB cases. Our study findings could contribute towards Brunei’s goal of achieving TB elimination by 2050, and could also contribute to the current limited scientific literature on EPTB epidemiology, particularly among Southeast Asian countries.

## Methods

### Data collection

This is a retrospective cohort study involving all registered TB cases (including PTB and EPTB) identified in Brunei Darussalam between January 2001 and December 2018. These cases were registered by the National Tuberculosis Coordination Centre (NTCC), a centre established in 2000 to implement, monitor, coordinate and evaluate TB prevention and control activities in the country [13]. All confirmed TB cases (initially presented at both public and private sector) are reported to NTCC or to its 3 directly observed treatment (DOTS) centres located in each of the 3 remaining districts for diagnosis, treatment and follow-up. The country’s population primarily resides near to the coastline, and that all 4 centres (NTCC and DOTS) were located near to major towns in each district (S1 Fig). The capital city is located at the Brunei-Muara district, the smallest district by land size but the most populated (where 72.3% of the country’s population resides [11]). Case information collected includes the year of diagnosis, sociodemographic details (age, gender, nationality, and district of residence), and details specific to the disease (type of TB, anatomic sites affected, mode of diagnosis and treatment outcome).

### Case definitions

A PTB case is defined as one with TB involving lung parenchyma, while a EPTB case is one with TB involving other organs except lungs [13]. The type of EPTB is defined based on the anatomical site where TB bacilli was found. In cases where TB bacilli was found in more than 1 anatomic site, the case was classified based on the clinical severity of the affected sites. Following the national guidelines, miliary TB (defined as cases with TB involving >1 organ including the lung) was classified as a PTB case. As such, it was not possible to classify cases as with concurrent PTB and EPTB.

We classified type of EPTB into 7 categories based on the main anatomical site affected: lymphatic, pleural, skeletal, gastrointestinal, genitourinary, meningeal, and other (consists of uncommon sites such as breast, skin, brain and eye). The mode of diagnosis was divided into 6 categories: tissue biopsy, cytology (test for fluid specimens collected via fine needle aspirate, endoscopy or bronchoalveolar lavage), radiology, clinical diagnosis alone, mycobacterium TB DNA (MTB DNA; a test for fluid specimens and performed if there was adequate growth in culture) and others (including those diagnosed overseas). Diagnosis of EPTB was based on the presence of TB bacilli on at least one specimen and/or histological evidence consistent with active EPTB. In cases where specimens could not be collected (such as for spine and brain TB), then radiology is the main diagnostic tool.

Treatment outcomes were classified into 6 categories [10,16]: (1) cured, defined as smear- or culture-positive TB patient at the start of treatment who was smear- or culture-negative in the last month of treatment; (2) completed, defined as TB patient who has completed treatment without failure but without record that shows smear- or culture-negative result in the last month of treatment; (3) failure, defined as TB patient whose smear or culture is positive on or after the fifth month of treatment; (4) died, defined as TB patient who died for any reason before or during treatment; (5) lost to follow-up, defined as TB patient who did not start treatment or whose treatment was interrupted for ≥2 consecutive months; or (6) not evaluated, defined as TB patient whose treatment outcome is unknown (including those who are still undergoing treatment) or who decided to seek treatment elsewhere, other than in Brunei. Not evaluated cases were mostly foreign workers who were diagnosed with active TB (either PTB or EPTB). In accordance with Brunei’s Immigration Act (Laws of Brunei, 2002), such workers would have their employment terminated and asked to return to their home country to continue their TB treatment.

### Statistical analysis

First, annual proportions of PTB and EPTB cases were calculated, using the total number of cases reported in each respective year as the denominator. Similarly, annual proportions of each EPTB type were calculated using the total number of EPTB cases reported in each respective year as the denominator. Second, socio-demographic characteristics of all groups (overall, PTB, overall EPTB and specific EPTB types) were described. Modes of diagnosis and treatment outcome were also described for overall EPTB and specific EPTB types. Third, group comparison tests were conducted using Chi-square or Fisher’s Exact, or Kruskal-Wallis tests, whichever appropriate. Fourth, logistic regression analysis was done to determine any factors associated with EPTB diagnosis. Variables with p-value of p<0.1 at the univariate analysis stage were included into the multiple logistic regression model, except for age-group and gender which were included *a priori*. Fifth, chi-square trend test was used to analyse trends between the proportion of cases by EPTB types. A p-value of <0.05 was considered statistically significant for all analyses. Microsoft Excel was used for data collection and result reporting (as tables and graphs). Statistical analyses were performed using R (ver. 3.6.3) [17].

### Ethics statement

Ethical approval was given by the Medical and Health Research and Ethics Committee, Ministry of Health, Brunei Darussalam (Ref: MHREC/UBD/2019/2). Patient informed consent was waived as data was retrospectively collected as part of the national TB surveillance, and only de-identified data was used in all analyses. Patients or the public were not involved in the design, or conduct, or reporting, or dissemination plans of our research.

### Patient and Public Involvement statement

Patients and/or public were not involved in the design, conduct, and result dissemination of this study.

## Results

A total of 3,916 TB cases were included in the study, out of which 19.0% (n=743) were diagnosed as EPTB. Among all TB cases (Table 1), their median age (interquartile range, IQR) was 44.0 years (IQR: 30.0 – 60.0), and mainly consisted of males (59.7%, n= 2338), locals (73.7%, n= 2886), and residing at Brunei-Muara district (63.7%, n= 2494). Among the EPTB cases, their median age (IQR) was 40.0 years (IQR: 27.5 – 56.0), about half were females (50.6%, n= 376), and mainly consisted of locals (78.6%, n= 582), and residing at Brunei-Muara district (70.6%, n= 525). When compared to PTB cases (Table 1), EPTB cases tend to be locals (adjusted Odds Ratio, aOR = 1.65 [95% Confidence Interval (95% CI): 1.35, 2.03]), and residing at Brunei-Muara district (Table 1).

**Table 1.** Characteristics of pulmonary TB (PTB) and extrapulmonary TB (EPTB) cases in Brunei, 2001-2018.

| Characteristics | Total (n = 3916) | Pulmonary TB (n = 3173) | Extrapulmonary TB (n = 743) | Crude OR | 95% CI | Adjusted OR* | 95% CI |
| --- | --- | --- | --- | --- | --- | --- | --- |
| **Age Group in years** |  |  |  |  |  |  |  |
| 0-24 | 551 (14.1) | 423 (13.3) | 128 (17.2) | **1.61** | **1.23, 2.12** | 1.4 | 0.96, 2.04 |
| 25-44 | 1448 (37.0) | 1152 (36.3) | 296 (39.8) | **1.37** | **1.09, 1.73** | 1.18 | 0.86, 1.62 |
| 45-64 | 1139 (29.1) | 943 (29.7) | 196 (26.4) | 1.11 | 0.87, 1.42 | 0.97 | 0.71, 1.34 |
| >65 | 778 (19.8) | 655 (20.7) | 123 (16.6) | 1 |  | 1 |  |
| **Gender** |  |  |  |  |  |  |  |
| Male | 2338 (59.7) | 1971 (62.2) | 367 (49.4) | 1 |  | 1 |  |
| Female | 1578 (40.3) | 1202 (37.8) | 376 (50.6) | **1.68** | **1.43,1.98** | 1.06 | 0.70,1.58 |
| **Nationality** |  |  |  |  |  |  |  |
| Local | 2886 (73.7) | 2304 (72.6) | 582 (78.3) | **1.36** | **1.13, 1.65** | **1.65** | **1.35,2.03** |
| Foreign | 1030 (26.3) | 869 (27.4) | 161 (21.7) | 1 |  | 1 |  |
| **District of residence** |  |  |  |  |  |  |  |
| Brunei Muara | 2494 (63.7) | 1969 (62.0) | 525 (70.6) | 1 |  | 1 |  |
| Belait | 736 (18.8) | 624 (19.7) | 112 (15.1) | **0.67** | **0.54,0.84** | **0.69** | **0.55,0.87** |
| Tutong | 540 (13.8) | 450 (14.2) | 90 (12.1) | **0.75** | **0.58,0.95** | **0.74** | **0.57,0.95** |
| Temburong | 146 (3.7) | 130 (4.1) | 16 (2.2) | **0.46** | **0.26,0.76** | **0.46** | **0.26,0.76** |
| *Model was also adjusted with interaction term (age group & gender). Significant positive interaction observed between gender & age group 25-44 years; Full model results with interaction term are shown in S1 Table. |  |  |  |  |  |  |  |

Among the 743 EPTB cases, the common type diagnosed was lymphatic (44.8%, n=333), followed by pleural (19.4%, n=144) and skeletal (11.6%, n=86; Figure 1). The male to female ratio varied slightly between EPTB types, with greater disproportion towards females (for lymphatic EPTB) and males (for pleural and meningeal EPTB; Table 2). The main modes of diagnosis were tissue biopsy (73.6% for overall EPTB) and radiologic assessment (18.3% for overall EPTB). High proportions of treatment completed (72.0%) or cured (7.7%) cases were also observed among all EPTB types. The overall mortality rate was 7.0% (n= 52), with disproportionately higher mortality for meningeal EPTB cases (6 out of 22 cases, 27.3%).

**Figure 1.**
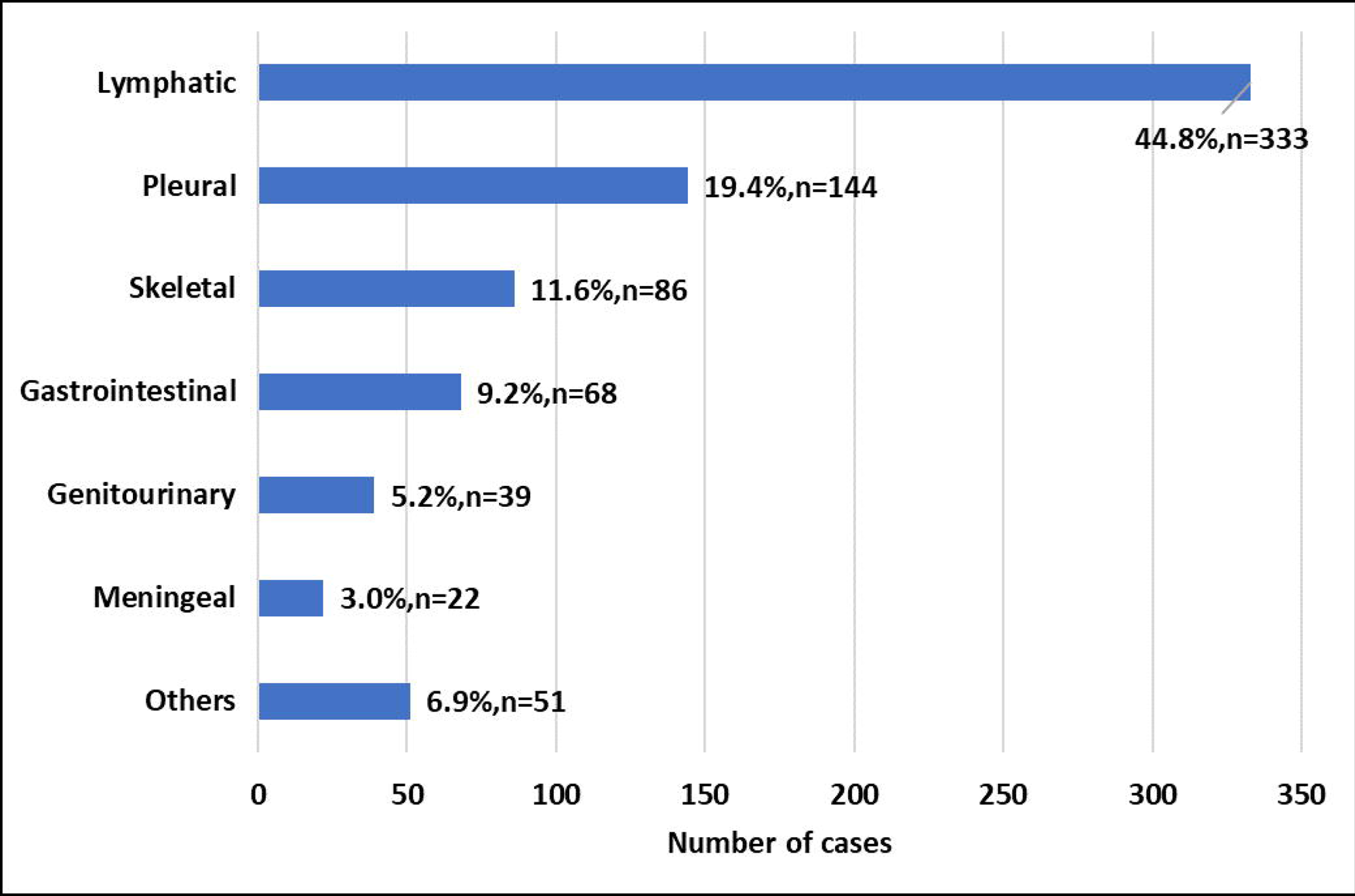
Types of extrapulmonary TB (EPTB) disease among 743 EPTB cases in Brunei, 2001-2018.

**Table 2.** Distribution of extrapulmonary TB (EPTB) by demographic and clinical characteristics, Brunei 2001-2018.

| Characteristic | Total<br>EPTB (n =<br>743) n(%) | Lymphatic<br>(n = 333)<br>n(%) | Pleural<br>(n = 144)<br>n(%) | Skeletal (n<br>= 86) n(%) | Gastrointestinal<br>(n = 68) n(%) | Genitourinary<br>(n = 39)<br>n(%) | Meningeal<br>(n = 22)<br>n(%) | Others (n<br>= 51)<br>n(%) |
| --- | --- | --- | --- | --- | --- | --- | --- | --- |
| **Median age in years (IQR)** | 40.0 (27.5 - 56.0) | 32.0 (24.0 - 45.0) | 46.0 (31.8 - 68.0) | 52.5 (34.8 - 64.0) | 45.0 (31.0 - 66.0) | 53.0 (41.0 - 60.5) | 41.0 (29.3 - 56.8) | 45.0 (33.5 - 53.5) |
| **Age range (years)** | 1 to 95 | 1 to 84 | 1 to 97 | 15 to 85 | 18 to 83 | 22 to 81 | 17 to 85 | 8 to 80 |
| **Age group (years)** |  |  |  |  |  |  |  |  |
| 0-4 | 7 (0.9) | 6 (1.8) | 1 (0.7) | 0 (0.0) | 0 (0.0) | 0 (0.0) | 0 (0.0) | 0 (0.0) |
| 5-24 | 122 (16.4) | 79 (23.7) | 17 (11.8) | 12 (14.0) | 3 (4.4) | 3 (7.7) | 3 (13.6) | 5 (9.8) |
| 25-44 | 296 (39.8) | 159 (47.7) | 49 (34.0) | 19 (22.1) | 30 (44.1) | 10 (25.6) | 10 (45.5) | 19 (37.3) |
| 45-64 | 196 (26.4) | 66 (19.8) | 36 (25.0) | 34 (39.5) | 15 (22.1) | 19 (48.7) | 4 (18.2) | 22 (43.1) |
| >65 | 123 (16.5) | 23 (6.9) | 41 (28.5) | 21 (24.4) | 20 (29.4) | 7 (18.0) | 5 (22.7) | 5 (9.8) |
| **Gender** |  |  |  |  |  |  |  |  |
| Male | 367 (49.4) | 126 (37.8) | 100 (69.4) | 40 (46.5) | 35 (51.5) | 22 (56.4) | 14 (63.6) | 30 (58.8) |
| Female | 376 (50.6) | 207 (62.2) | 44 (30.6) | 46 (53.5) | 33 (48.5) | 17 (43.6) | 8 (36.4) | 21 (41.2) |
| **Nationality** |  |  |  |  |  |  |  |  |
| Local | 582 (78.3) | 237 (71.2) | 109 (75.7) | 80 (93.0) | 61 (89.7) | 33 (84.6) | 17 (77.3) | 45 (88.2) |
| Non-Local | 161 (21.7) | 96 (28.8) | 35 (24.3) | 6 (7.0) | 7 (10.3) | 6 (15.4) | 5 (22.7) | 6 (11.8) |
| **District** |  |  |  |  |  |  |  |  |
| Brunei-Muara | 525 (70.6) | 246 (73.9) | 102 (70.8) | 56 (65.1) | 45 (66.2) | 28 (71.8) | 13 (59.1) | 35 (68.6) |
| Belait | 112 (15.1) | 47 (14.1) | 29 (20.1) | 10 (11.6) | 10 (14.7) | 5 (12.8) | 3 (13.6) | 8 (15.7) |
| Tutong | 90 (12.1) | 38 (11.4) | 10 (7.0) | 15 (17.5) | 12 (17.6) | 4 (10.3) | 6 (27.3) | 5 (9.8) |
| Temburong | 16 (2.2) | 2 (0.6) | 3 (2.1) | 5 (5.8) | 1 (1.5) | 2 (5.1) | 0 (0.0) | 3 (5.9) |
| **Mode of diagnosis** |  |  |  |  |  |  |  |  |
| Tissue biopsy | 547 (73.6) | 261 (78.4) | 91 (63.2) | 63 (73.2) | 51 (75.0) | 31 (79.5) | 17 (77.3) | 33 (64.7) |
| Clinical alone | 2 (0.3) | 1 (0.3) | 1 (0.7) | 0 (0.0) | 0 (0.0) | 0 (0.0) | 0 (0.0) | 0 (0.0) |
| Cytology | 15 (2.0) | 0 (0.0) | 12 (8.3) | 0 (0.0) | 3 (4.4) | 0 (0.0) | 0 (0.0) | 0 (0.0) |
| MTB-DNA | 30 (4.0) | 9 (2.7) | 2 (1.4) | 5 (5.8) | 3 (4.4) | 2 (5.1) | 4 (18.2) | 5 (9.8) |
| Others | 13 (1.8) | 3 (0.9) | 4 (2.8) | 4 (4.7) | 0 (0.0) | 1 (2.6) | 0 (0.0) | 1 (2.0) |
| Radiology | 136 (18.3) | 59 (17.7) | 34 (23.6) | 12 (16.3) | 11 (16.2) | 5 (12.8) | 1 (4.5) | 12 (23.5) |
| **Outcome of treatment** |  |  |  |  |  |  |  |  |
| Cured | 57 (7.7) | 29 (8.7) | 11 (7.7) | 6 (7.0) | 4 (5.8) | 2 (5.1) | 1 (4.5) | 4 (7.8) |
| Completed | 535 (72.0) | 235 (70.6) | 93 (65.0) | 67 (77.9) | 56 (82.4) | 35 (89.7) | 10 (45.5) | 39 (76.5) |
| Died | 52 (7.0) | 14 (4.2) | 16 (11.2) | 8 (9.3) | 5 (7.4) | 1 (2.6) | 6 (27.3) | 2 (3.9) |
| Lost to follow-up | 3 (0.4) | 1 (0.3) | 0 (0.0) | 1 (1.2) | 1 (1.5) | 0 (0.0) | 0 (0.0) | 0 (0.0) |
| Treatment failure | 1 (0.1) | 0 (0.0) | 0 (0.0) | 0 (0.0) | 0 (0.0) | 0 (0.0) | 0 (0.0) | 1 (2.0) |
| Not evaluated | 95 (12.8) | 54 (16.2) | 23 (16.0) | 4 (4.6) | 2 (2.9) | 1 (2.6) | 5 (22.7) | 5 (9.8) |

IQR: Interquartile range; MTB-DNA: Mycobacterium TB DNA
S2 Table shows the side -by-side comparison with PTB case counts & group comparison results between specific EPTB types

Multiple logistic regression analysis (Table 3) showed that age is associated with specific EPTB types: adjusted odds of lymphatic EPTB affecting younger people were at least 1.94 times higher when compared to those >65 years, and those aged between 25-64 years had at least 3.15 times more odds to develop EPTB in other sites. On the other hand, younger people had lower adjusted odds of developing pleural (aOR for 25-44: 0.54 [95% CI: 0.32, 0.89] and aOR for 45-64: 0.42 [95% CI: 0.24, 0.73]) and gastrointestinal EPTB (aOR for 0-24: 0.26 [95% CI: 0.06, 0.78]), when compared to those >65 years. Females had higher adjusted odds of developing lymphatic (aOR: 2.45 [95% CI: 1.94, 3.11]) and skeletal (aOR: 1.91 [95% CI: 1.24, 2.95]) EPTB but had lower adjusted odds of developing pleural EPTB (aOR: 0.41 [95% CI: 0.17, 0.90]) when compared to males. Also, locals had higher adjusted odds of developing skeletal (aOR: 4.44 [95% CI: 2.04, 11.69]), gastrointestinal (aOR: 3.91 [95% CI: 1.84, 9.66]) and other types of EPTB (aOR: 3.42 [95% CI: 1.53, 9.14]) when compared to foreign residents. Lastly, residents of Brunei-Muara district have higher odds of developing lymphatic and pleural EPTB when compared to other smaller districts.

**Table 3.**
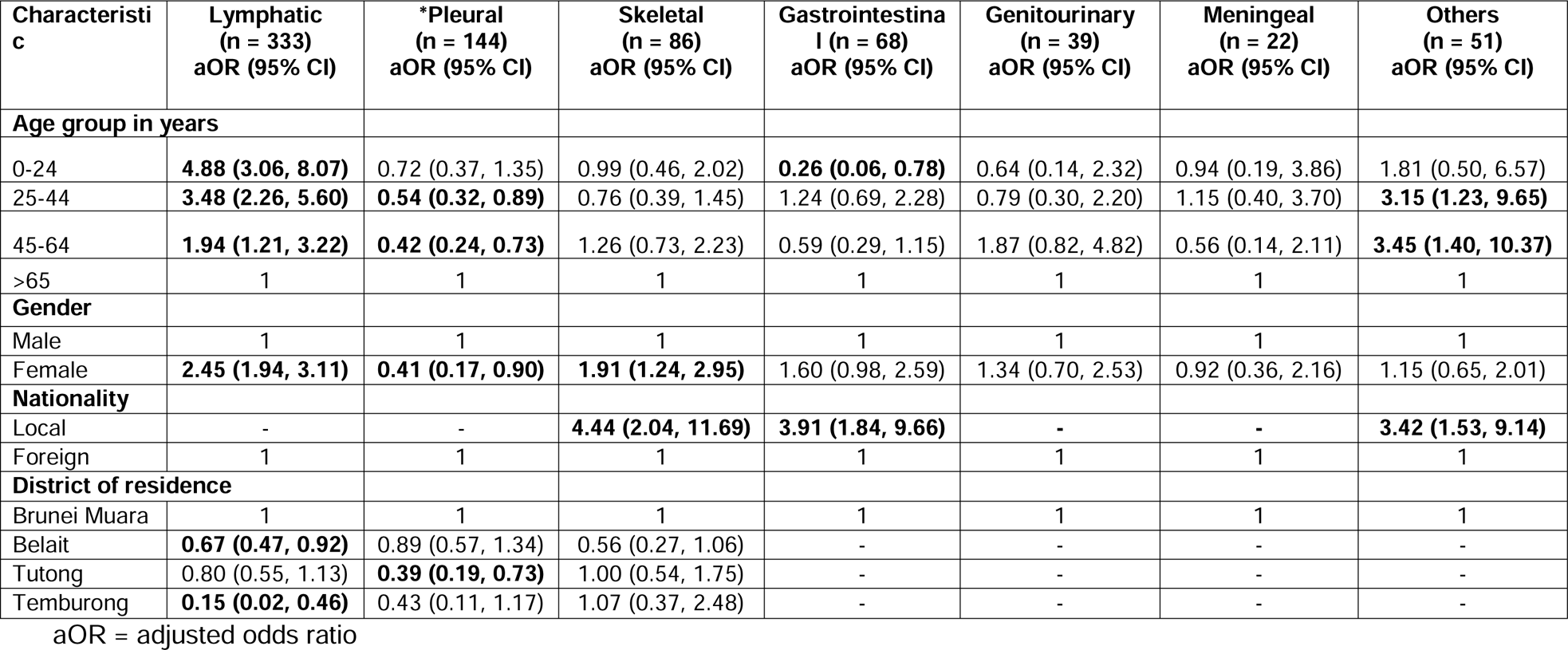
Multiple logistic regression results for 7 types of extrapulmonary TB (EPTB) compared with pulmonary TB (PTB), Brunei 2001-2018.

We analysed trends by types of EPTB during the 18-year study period. The proportions of both PTB and EPTB cases remained steady with no significant trend differences (Figure 2), where EPTB cases accounted between 12.1% and 24.4% of the annual TB cases. Chi-square trend tests revealed no significant trend differences for all EPTB types, when compared with PTB cases (Figure 2).

**Figure 2.**
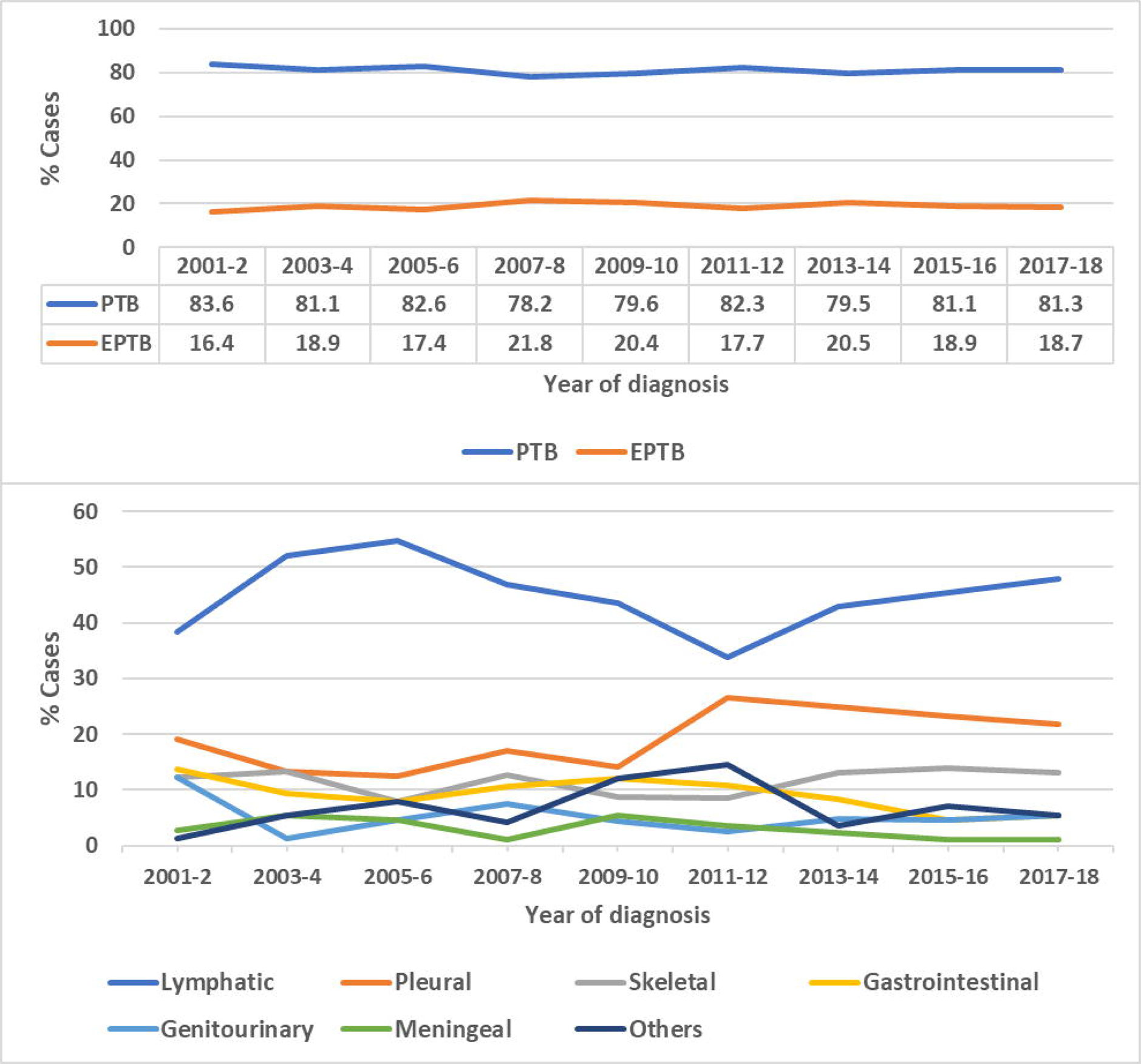
Relative proportion of (top) pulmonary TB (PTB) and extrapulmonary TB (EPTB) cases, and (bottom) of all 7 types extrapulmonary TB (EPTB) by year of diagnosis (reported in 2-year intervals).

## Discussion

In this 18-year retrospective analysis, we found that about one-fifth of all TB cases in Brunei (19.0%) were diagnosed as EPTB. This finding is line with that previously reported in population-based studies from other regions [3–6,18–20], which ranges between 10.4% and 21.8% but lower than that reported from England and Wales (41%) [21]. We also found that lymphatic EPTB was the most dominant type in Brunei, a finding similar to countries with low [19,21], intermediate [18], and high [8] TB-burden. Differences in the dominant EPTB types can be seen in other studies particularly from high TB-burden countries, such as disseminated EPTB in Ghana [20], skeletal [7] and pleural [4] EPTB in China. This point further adds on to the known observation that dominant EPTB types can vary across different geographical areas, possibly due (but not limited) to differences in socio-demographics, diagnostic capability, and access to health care.

In addition, we observed interesting age and gender differences in the association between specific EPTB types. On one hand, younger age groups were associated with lymphatic and other EPTB while female gender was associated with lymphatic, skeletal and gastrointestinal EPTB. Both factors (younger age and females) have previously been reported as factors of developing EPTB in high, intermediate and low TB-burden countries [5,7–9,18,19]. On the other hand, we found that pleural EPTB was associated with males and older age; a finding consistent with another study in a Chinese city [4], but contrasts with another study of Chinese inpatients where pleural EPTB was associated with males and younger age [7]. Taken together, we believe that factors associated with EPTB is more dependent on the specific type of EPTB as the main anatomic site. This implies that it would be more informative to analyse the factors associated with specific types EPTB, rather than for overall EPTB cases. Having said that, there is also a need to disentangle the presence of multi-site TB infections, with or without lung involvement. Recent studies have reported that concurrent EPTB with other EPTB types or PTB was common among EPTB inpatients [22], and that the degree of association between socio-demographic factors (age and gender) and the different types of concurrent EPTB tends to vary [23].

We found that local Bruneian residents had at least 3 times higher odds of developing skeletal, gastrointestinal, and other EPTB, albeit less precise due to the quite wide 95% confidence intervals. This is in contrast with previous studies from low TB-burden countries (United States [19] and European Union [6]) that reported foreign residents as a factor associated with EPTB. A plausible explanation is that upon initial suspicion of EPTB diagnosis, foreign workers may opt to refuse further diagnostic tests and be sent back to home country, due to the monetary costs involved. Foreign residents makes up 28% of Brunei’s labour force, with significant proportion employed as low-skilled workers in construction, wholesale and retail trade [24,25]. Foreign workers are required to undergo medical check-up upon start and renewal of their employment contract, and those diagnosed with active TB (PTB or EPTB) would be asked to return to their home country for treatment [26]. Employers could decide it is more cost-effective to send their foreign workers home upon initial suspicion of EPTB diagnosis during medical check-up, particularly in EPTB types that are difficult to diagnose and require follow-up testing, such as skeletal EPTB. While TB testing and treatment is provided free of charge for everyone, foreign nationals are charged fees for diagnostic tests conducted prior to EPTB diagnosis confirmation; this is free for locals. This could be one reason why locals instead made up the majority of all EPTB cases in this study, and that 90.5% (86/95) of EPTB cases whose treatment outcome was classified as “Not evaluated” were foreign residents.

Lastly, we observed those living in Brunei-Muara district were positively associated with EPTB (for overall, lymphatic and pleural). One explanation relates to population density; Brunei-Muara is the smallest but most populated district in Brunei, where 72.3% of the country’s population resides, and where the capital city is located [11]. This district is also comparatively more urbanised; residing in urban areas was reported as one factor associated with EPTB in other countries [4,5,18]. While it is true that the capital city has more laboratory diagnostic facilities, assess to testing would not be an issue as clinical specimens are routinely sent from other districts. Similarly, there would also be no significant differences in terms of initially suspecting EPTB among clinicians across all districts even though the capital city has more diagnostic facilities.

It should be noted that access to healthcare is not a major issue in Brunei, as all 4 districts have their own DOTS centres and district hospitals to enable adequate patient follow-up and diagnostic testing. This is quite evident with the high treatment success rate observed in our study population, where 79.7% of all EPTB cases were either cured or completed their treatment. Our treatment success rate is slightly better than that reported in Malaysia (67.6%) [18] and Ghana (70.1%) [20]. Efforts were also made to confirm EPTB diagnosis; 73.6% of all EPTB cases in our study population were diagnosed through tissue biopsy and only 0.3% was diagnosed through clinical judgement. These observations suggest that the national TB programme is doing well in terms of EPTB case diagnosis, follow-up and treatment. Despite this, however, no trend changes could be observed for EPTB (also for PTB) cases during the study period. This finding suggests that the known stagnant situation for the overall TB incidence in Brunei since 2004 [13] is extended to both PTB and EPTB. One plausible explanation could be that people are simply unaware one can get TB in many parts of the body other than the lungs, and that it could manifest in the form of non-painful bumps; early symptoms that could be ignored and only seek medical attention when it becomes bigger or painful. Such delayed health seeking behaviour could thus result in the accumulation of EPTB cases over time. Promoting public awareness of EPTB together and with the same rigour as for PTB, using existing mechanisms already in place for health information dissemination, would be a useful and feasible step forward.

This study has several limitations. First, data entry errors are possible due to the study’s retrospective nature, and in some cases, it is not possible to confirm misdiagnosis or to clarify uncertainties in the data. However, efforts were made to review any suspected data entry error by re-checking from either hardcopy or electronic health records (EHR). Second, we were unable to distinguish between PTB cases with only lung involvement and those with miliary TB and/or concurrent PTB and EPTB. This is a consequence of defining all TB cases involving the lung as PTB (under the national TB guidelines), and suggests that our odds ratio estimates could be biased towards the null. Third, we were unable to collect data on other known factors associated with EPTB (such as low immune status, diabetic and HIV positive), as there were not available for the whole study period. This is because data were mainly collected through hardcopy records (with variable degree of completion) until 2013, when a national EHR system was established. However, this limitation may not greatly affect our results due to potentially small case numbers with co-morbidities. Using data from 2013 to 2018 (shorter but overlapping with this study’s time period), we have previously reported that 20.1% (53/264) of all EPTB cases were also diagnosed with diabetes [10]. Also, annual HIV incidence in Brunei is low; the annual number of HIV case notifications from 2014 to 2018 ranged between 13 and 41 cases [14]. Leveraging the EHR system already in place, co-morbidity information could be added the data collection checklist for TB cases and digitally recorded for future epidemiological studies. Despite these limitations, the strength of the study is the generalisability of the results to the Bruneian population, as all EPTB notifications in the country were captured in our dataset.

In conclusion, we observed 19.0% of all active TB cases during the 18-year study period had EPTB. Lymphatic EPTB was the most common type, followed by pleural and skeletal EPTB. We also observed that associations with specific EPTB types varies with age-group and gender. And that examining EPTB cases by their specific anatomical site would provide more information on risk factors. Including EPTB content into existing health information dissemination programmes would help raise public awareness on the condition, and in turn promotes early health seeking behaviour and early EPTB diagnosis. Future studies could be done to analyse the association between EPTB and presence of co-morbidities.

## Acknowledgements

We are grateful for all staff involved in data collection and reviewing process at NTCC and DOTS clinics.

## Data availability statement

The dataset used in the analysis is provided as Supplementary Information.

## Author contributions

LC, LMS and KT conceived the study.

RAH, and KT collected the data.

RAH, KT and LC supervised the study.

LC and LMS analysed the data and drafted the manuscript.

All authors read, revised and approved the manuscript.

## References

1 World Health Organization. Global Tuberculosis Report 2020. 2020. https://www.who.int/publications/i/item/9789240013131 (accessed 22 Dec 2020).

2 Houda Ben A, Makram K, Chakib M, et al. Extrapulmonary Tuberculosis: Update on the Epidemiology, Risk Factors and Prevention Strategies. Int J Trop Dis 2018;1:1–6. doi:10.23937/ijtd-2017/1710006

3 Lee JY. Diagnosis and Treatment of Extrapulmonary Tuberculosis. Tuberc Respir Dis (Seoul) 2015;78:47–55. doi:https://doi.org/10.4046/trd.2015.78.2.47

4 Wang X, Yang Z, Fu Y, et al. Insight to the epidemiology and risk factors of extrapulmonary tuberculosis in Tianjin, China during 2006-2011. PLoS One 2014;9. doi:10.1371/journal.pone.0112213

5 Gomes T, Reis-santos B, Bertolde A, et al. Epidemiology of extrapulmonary tuberculosis in BrazillJ: a hierarchical model. BMC Infect Dis 2014;14. doi:https://doi.org/10.1186/1471-2334-14-9

6 Sandgren A, Hollo V, van der Werf MJ. Extrapulmonary tuberculosis in the European union and European economic area, 2002 to 2011. Eurosurveillance 2013;18:1–9. doi:10.2807/ese.18.12.20431-en

7 Pang Y, An J, Shu W, et al. Epidemiology of extrapulmonary tuberculosis among inpatients, China, 2008-2017. Emerg Infect Dis 2019;25:457–64. doi:10.3201/eid2503.180572

8 Arega B, Mersha A, Minda A, et al. Epidemiology and the diagnostic challenge of extra-pulmonary tuberculosis in a teaching hospital in Ethiopia. PLoS One 2020;15:1–15. doi:10.1371/journal.pone.0243945

9 Yang Z, Kong Y, Wilson F, et al. Identification of Risk Factors for Extrapulmonary Tuberculosis. 2004.

10 Omar N, Wong J, Thu K, et al. Prevalence and associated factors of diabetes mellitus among tuberculosis patients in Brunei Darussalam: A 6-year retrospective cohort study. Int J Infect Dis 2021;105:267–73. doi:10.1016/j.ijid.2021.02.064

11 Department of Economic Planning and Development. Population and Housing Census. 2021.http://www.deps.gov.bn/SitePages/Population.aspx (accessed 15 May 2023).

12 World Health Organization. Brunei Darussalam Tuberculosis Country Profile. 2022. https://www.aidsdatahub.org/resource/brunei-darussalam-tuberculosis-country-profile-2022

13 Ministry of Health Brunei Darussalam. Guidelines for Tuberculosis Control in Brunei Darussalam. 2013. http://www.moh.gov.bn/SitePages/Downloads.aspx

14 World Health Organization. Country factsheet: Public health data at a glance - Brunei Darussalam. 2020.https://www.who.int/docs/default-source/wpro---documents/countries/brunei-darussalam/fact-sheet-brunei-darussalam.pdf?sfvrsn=b91eddf4_5. (accessed 1 Apr 2022).

15 The World Bank. Diabetes prevalence (% of population ages 20 to 79) - Brunei Darussalam. 2021.https://data.worldbank.org/indicator/SH.STA.DIAB.ZS?locations=BN (accessed 19 May 2023).

16 World Health Organization. Definitions and reporting framework for tuberculosis 2013 revision (updated December 2014). 2013;2019.https://www.who.int/tb/publications/definitions/en/

17 R Core Team. R: A language and environment for statistical computing. 2021.http://www.r-project.org/

18 Khan AH, Sulaiman SAS, Laghari M, et al. Treatment outcomes and risk factors of extra-pulmonary tuberculosis in patients with co-morbidities. BMC Infect Dis 2019;19. doi:10.1186/s12879-019-4312-9

19 Peto HM, Pratt RH, Harrington TA, et al. Epidemiology of Extrapulmonary Tuberculosis in the United States, 1993 – 2006. 2009;30333:1350–7. doi:10.1086/605559

20 Ohene S-A, Bakker MI, Ojo J, et al. Extra-pulmonary tuberculosis: A retrospective study of patients in Accra, Ghana. PLoS One 2019;14:e0209650. doi:10.1371/journal.pone.0209650

21 Kruijshaar ME, Abubakar I. Increase in extrapulmonary tuberculosis in England and Wales 1999 – 2006. Thorax 2009;:1090–5. doi:10.1136/thx.2009.118133

22 Kang W, Yu J, Du J, et al. The epidemiology of extrapulmonary tuberculosis in China: A large-scale multicenter observational study. PLoS One 2020;15:1–15. doi:10.1371/journal.pone.0237753

23 Kang W, Liu S, Du J, et al. Epidemiology of concurrent extrapulmonary tuberculosis in inpatients with extrapulmonary tuberculosis lesions in China: a large-scale observational multi-centre investigation. Int J Infect Dis 2022;115:79–85. doi:10.1016/j.ijid.2021.11.019

24 Department of Economic Planning and Statistics Ministry of Finance and Economy Brunei Darussalam. Labour Force Survey 2020. Bandar Seri Begawan: 2020. http://www.deps.gov.bn/SitePages/LabourForce.aspx

25 Chaw L, Abdul Hamid R, Koh KS, et al. Contact investigation of tuberculosis in Brunei Darussalam: Evaluation and risk factor analysis. BMJ Open Respir Res 2022;9:e001224. doi:10.1136/bmjresp-2022-001224

26 Laws of Brunei. Chapter 17 Immigration. 2002.https://www.ilo.org/wcmsp5/groups/public/@ed_protect/@protrav/@ilo_aids/documents/legaldocument/wcms_117279.pdf (accessed 12 Jan 2021).

