## Supplementary material for "Epidemiology of Extrapulmonary Tuberculosis in Brunei Darussalam: A retrospective cohort study": S1 Fig, S1 Table, S2 Table

S1 Fig. Topographic map of Brunei Darussalam showing the locations of NTCC near to Bandar Seri Begawan (Brunei's capital city in Brunei-Muara district), and the 3 DOTS (Directly Observed Therapy) clinics located near the major towns of each district (from left to right: Belait, Tutong, Brunei-Muara, and Temburong).

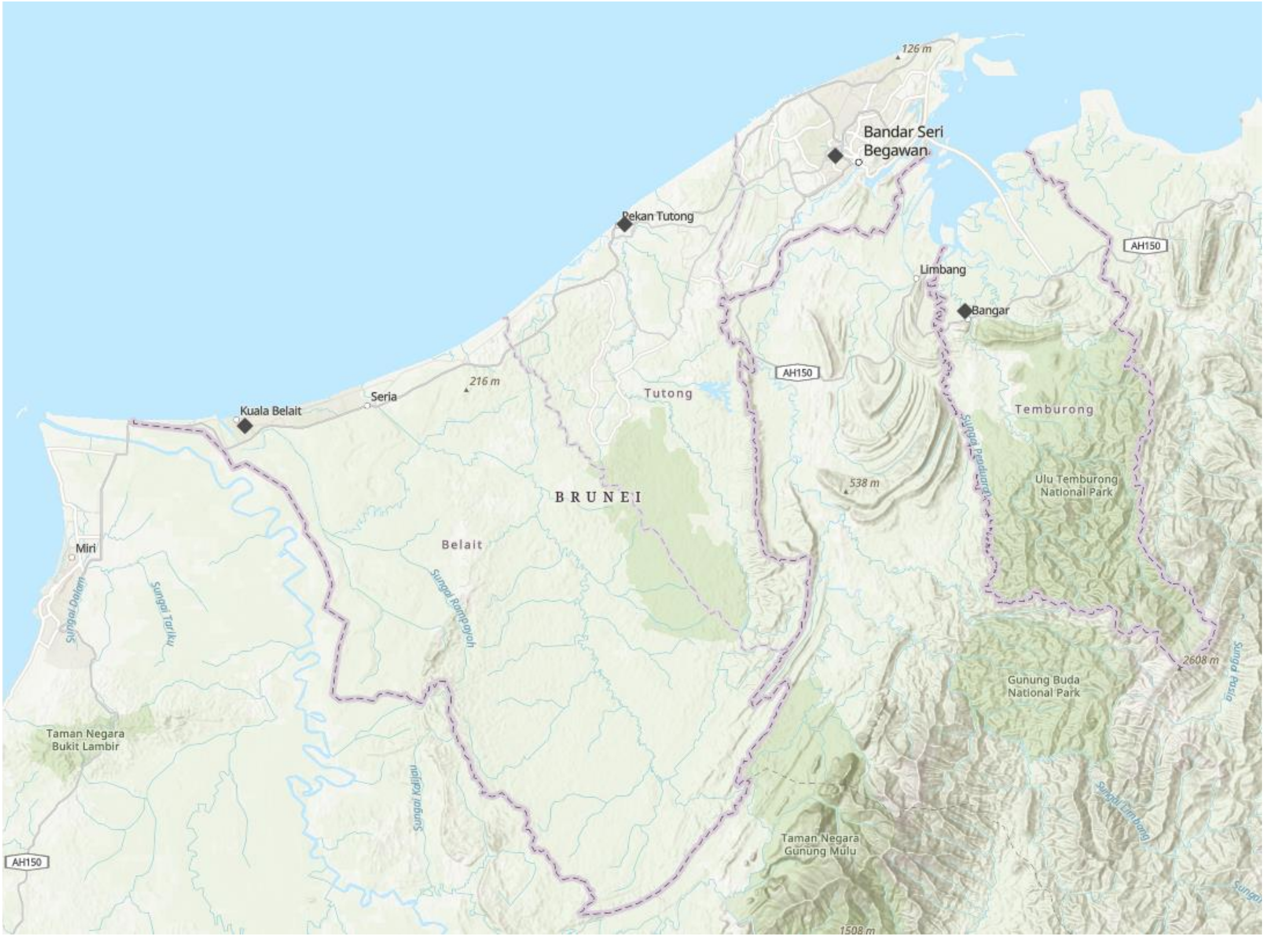


(Map created by author using ArcGIS online website)

S1 Table. Characteristics of pulmonary TB (PTB) and extrapulmonary TB (EPTB) cases in Brunei, 2001-2021: Complete logistic regression results (with interaction term)

| Characteristics | Total (n = 3916) | Pulmonary TB (n = 3173) | Extrapulmonary TB (n = 743) | Crude OR | 95% CI | Adjusted OR* | 95% CI |
|---|---|---|---|---|---|---|---|
| **Age Group in years** | | | | | | | |
| 0-24 | 551 (14.1) | 423 (13.3) | 128 (17.2) | **1.61** | **1.23, 2.12** | 1.4 | 0.96, 2.04 |
| 25-44 | 1448 (37.0) | 1152 (36.3) | 296 (39.8) | **1.37** | **1.09, 1.73** | 1.18 | 0.86, 1.62 |
| 45-64 | 1139 (29.1) | 943 (29.7) | 196 (26.4) | 1.11 | 0.87, 1.42 | 0.97 | 0.71, 1.34 |
| >65 | 778 (19.8) | 655 (20.7) | 123 (16.6) | 1 | | 1 | |
| **Gender** | | | | | | | |
| Male | 2338 (59.7) | 1971 (62.2) | 367 (49.4) | 1 | | 1 | |
| Female | 1578 (40.3) | 1202 (37.8) | 376 (50.6) | **1.68** | **1.43,1.98** | 1.06 | 0.70,1.58 |
| **Nationality** | | | | | | | |
| Local | 2886 (73.7) | 2304 (72.6) | 582 (78.3) | **1.36** | **1.13, 1.65** | **1.65** | **1.35,2.04** |
| Non-Local | 1030 (26.3) | 869 (27.4) | 161 (21.7) | 1 | | 1 | |
| **District of residence** | | | | | | | |
| Brunei Muara | 2494 (63.7) | 1969 (62.0) | 525 (70.6) | 1 | | 1 | |
| Belait | 736 (18.8) | 624 (19.7) | 112 (15.1) | **0.67** | **0.54,0.84** | **0.69** | **0.55,0.87** |
| Tutong | 540 (13.8) | 450 (14.2) | 90 (12.1) | **0.75** | **0.58.0.95** | **0.74** | **0.57,0.95** |
| Temburong | 146 (3.7) | 130 (4.1) | 16 (2.2) | **0.46** | **0.26,0.76** | **0.46** | **0.26,0.76** |
| **Interaction term** | | | | | | | |
| Age group (1-24) & Female gender | | | | | | 1.40 | 0.79,2.49 |
| Age group (25-44) & Female gender | | | | | | **1.83** | **1.13,2.98** |
| Age group (45-64) & Female gender | | | | | | 1.54 | 0.92,2.59 |

S2 Table. Distribution of extrapulmonary TB (EPTB) by demographic and clinical characteristics, Brunei 2001-2018: Version with PTB statistics and group comparison results between specific EPTB types

| **Characteristic** | **Total PTB (n=3173) n(%)** | **Total EPTB (n = 743) n(%)** | **Lymphatic (n = 333) n(%)** | **Pleural (n = 144) n(%)** | **Skeletal (n = 86) n(%)** | **Gastrointestinal (n = 68) n(%)** | **Genitourinary (n = 39) n(%)** | **Meningeal (n = 22) n(%)** | **Others (n = 51) n(%)** | **p value^** |
|---|---|---|---|---|---|---|---|---|---|---|
| **Median age in years (IQR)** | 45.0 (31.0-61.0) | 40.0 (27.5 - 56.0) | 32.0 (24.0 - 45.0) | 46.0 (31.8 - 68.0) | 52.5 (34.8 - 64.0) | 45.0 (31.0 - 66.0) | 53.0 (41.0 - 60.5) | 41.0 (29.3 - 56.8) | 45.0 (33.5 - 53.5) | <0.001* |
| **Age range (years)** | 1 to 97 | 1 to 95 | 1 to 84 | 1 to 97 | 15 to 85 | 18 to 83 | 22 to 81 | 17 to 85 | 8 to 80 | |
| **Age group (years)** | | | | | | | | | | |
| 0-4 | 4 (0.1) | 7 (0.9) | 6 (1.8) | 1 (0.7) | 0 (0.0) | 0 (0.0) | 0 (0.0) | 0 (0.0) | 0 (0.0) | <0.001 |
| 5-24 | 419 (13.2) | 122 (16.4) | 79 (23.7) | 17 (11.8) | 12 (14.0) | 3 (4.4) | 3 (7.7) | 3 (13.6) | 5 (9.8) | |
| 25-44 | 1152 (36.3) | 296 (39.8) | 159 (47.7) | 49 (34.0) | 19 (22.1) | 30 (44.1) | 10 (25.6) | 10 (45.5) | 19 (37.3) | |
| 45-64 | 943 (29.7) | 196 (26.4) | 66 (19.8) | 36 (25.0) | 34 (39.5) | 15 (22.1) | 19 (48.7) | 4 (18.2) | 22 (43.1) | |
| >65 | 655 (20.7) | 123 (16.5) | 23 (6.9) | 41 (28.5) | 21 (24.4) | 20 (29.4) | 7 (18.0) | 5 (22.7) | 5 (9.8) | |
| **Gender** | | | | | | | | | | |
| Male | 1971 (62.2) | 367 (49.4) | 126 (37.8) | 100 (69.4) | 40 (46.5) | 35 (51.5) | 22 (56.4) | 14 (63.6) | 30 (58.8) | <0.001 |
| Female | 1202 (37.8) | 376 (50.6) | 207 (62.2) | 44 (30.6) | 46 (53.5) | 33 (48.5) | 17 (43.6) | 8 (36.4) | 21 (41.2) | |
| **Nationality** | | | | | | | | | | |
| Local | 2304 (72.6) | 582 (78.3) | 237 (71.2) | 109 (75.7) | 80 (93.0) | 61 (89.7) | 33 (84.6) | 17 (77.3) | 45 (88.2) | <0.001 |
| Non-Local | 869 (27.4) | 161 (21.7) | 96 (28.8) | 35 (24.3) | 6 (7.0) | 7 (10.3) | 6 (15.4) | 5 (22.7) | 6 (11.8) | |
| **District** | | | | | | | | | | |
| Brunei-Muara | 1969 (62.0) | 525 (70.6) | 246 (73.9) | 102 (70.8) | 56 (65.1) | 45 (66.2) | 28 (71.8) | 13 (59.1) | 35 (68.6) | 0.02 |
| Belait | 624 (19.7) | 112 (15.1) | 47 (14.1) | 29 (20.1) | 10 (11.6) | 10 (14.7) | 5 (12.8) | 3 (13.6) | 8 (15.7) | |
| Tutong | 450 (14.2) | 90 (12.1) | 38 (11.4) | 10 (7.0) | 15 (17.5) | 12 (17.6) | 4 (10.3) | 6 (27.3) | 5 (9.8) | |
| Temburong | 130 (4.1) | 16 (2.2) | 2 (0.6) | 3 (2.1) | 5 (5.8) | 1 (1.5) | 2 (5.1) | 0 (0.0) | 3 (5.9) | |
| **Mode of diagnosis** | | | | | | | | | | |
| Tissue biopsy | 53 (1.7) | 547 (73.6) | 261 (78.4) | 91 (63.2) | 63 (73.2) | 51 (75.0) | 31 (79.5) | 17 (77.3) | 33 (64.7) | <0.001 |
| Clinical alone | 150 (4.7) | 2 (0.3) | 1 (0.3) | 1 (0.7) | 0 (0.0) | 0 (0.0) | 0 (0.0) | 0 (0.0) | 0 (0.0) | |
| Cytology | 39 (1.2) | 15 (2.0) | 0 (0.0) | 12 (8.3) | 0 (0.0) | 3 (4.4) | 0 (0.0) | 0 (0.0) | 0 (0.0) | |
| MTB-DNA | 13 (0.4) | 30 (4.0) | 9 (2.7) | 2 (1.4) | 5 (5.8) | 3 (4.4) | 2 (5.1) | 4 (18.2) | 5 (9.8) | |

| | | | | | | | | | | |
|---|---|---|---|---|---|---|---|---|---|---|
| Others | 9 (0.3) | 13 (1.8) | 3 (0.9) | 4 (2.8) | 4 (4.7) | 0 (0.0) | 1 (2.6) | 0 (0.0) | 1 (2.0) | |
| Radiology | 183 (5.8) | 136 (18.3) | 59 (17.7) | 34 (23.6) | 12 (16.3) | 11 (16.2) | 5 (12.8) | 1 (4.5) | 12 (23.5) | |
| Acid-fast bacilli smear/culture | 2726 (85.9) | 0 (0.0) | | | | | | | | |
| **Outcome of treatment** | | | | | | | | | | |
| Cured | 1660 (52.3) | 57 (7.7) | 29 (8.7) | 11 (7.7) | 6 (7.0) | 4 (5.8) | 2 (5.1) | 1 (4.5) | 4 (7.8) | |
| Completed | 559 (17.6) | 535 (72.0) | 235 (70.6) | 93 (65.0) | 67 (77.9) | 56 (82.4) | 35 (89.7) | 10 (45.5) | 39 (76.5) | |
| Died | 243 (7.7) | 52 (7.0) | 14 (4.2) | 16 (11.2) | 8 (9.3) | 5 (7.4) | 1 (2.6) | 6 (27.3) | 2 (3.9) | |
| Lost to follow-up | 26 (0.8) | 3 (0.4) | 1 (0.3) | 0 (0.0) | 1 (1.2) | 1 (1.5) | 0 (0.0) | 0 (0.0) | 0 (0.0) | <0.001 |
| Treatment failure | 3 (0.1) | 1 (0.1) | 0 (0.0) | 0 (0.0) | 0 (0.0) | 0 (0.0) | 0 (0.0) | 0 (0.0) | 1 (2.0) | |
| Not evaluated | 682 (21.5) | 95 (12.8) | 54 (16.2) | 23 (16.0) | 4 (4.6) | 2 (2.9) | 1 (2.6) | 5 (22.7) | 5 (9.8) | |

^Group comparison analyses were conducted between the 7 specific EPTB types

*Post-hoc dunn test revealed that age of lymphatic EPTB patients were significantly different than all other EPTB types (except for meningeal)

IQR: Interquartile range; MTB-DNA: Mycobacterium TB DNA
